# Neuroimaging and behavioural biomarkers of post-stroke cognitive recovery outcomes

**DOI:** 10.64898/2026.05.12.26353056

**Authors:** Margaret Jane Moore, Stephanie J. Forkel, Nele Demeyere

## Abstract

Lesion anatomy has been widely used to study post stroke cognitive outcomes, but it is unclear whether lesion-based measures provide clinically meaningful prognostic information beyond established predictors. Stroke survivors (n = 408) completed the Oxford Cognitive Screen (OCS) during acute hospitalisation and at chronic (6-month) follow-up. Lesion characteristics and structural disconnection profiles associated with chronic OCS scores were identified using ROI-level, voxel-level and structural network disconnection lesion mapping approaches. The incremental predictive value of these measures, relative to acute behaviour and pre-morbid brain health, was evaluated using regression analyses, receiver operating curve (ROC) and support vector regression (SVR) models predicting continuous chronic scores.

Significant lesion and disconnection correlates of chronic cognitive impairment were identified for 9/10 OCS subtests. The extent of damage to these correlates was significantly associated with chronic cognitive scores, but their diagnostic utility for identifying persistent impairment was low under conventional thresholds (AUC mean = 0.59, range= 0.46–0.66). Acute cognitive task performance was the single best predictor of chronic cognition (AUC mean = 0.66, range = 0.41–0.95). In multivariate analyses, SVR models trained on acute cognitive performance and regional atrophy severity scores both outperformed models trained on lesion anatomy or structural disconnection across most cognitive domains. SVR models combining anatomical, disconnection and behavioural predictors did not improve predictions accuracy relative to behaviour- or atrophy-only models.

Together, these findings demonstrate that statistically significant lesion–outcome relationships do not necessarily translate into clinically useful prognostic indicators. In a large, clinically representative stroke cohort, detailed lesion-based measures provided limited incremental prognostic value beyond acute cognitive assessment and coarse brain health markers. These results highlight the importance of explicitly evaluating predictive utility when developing prognostic models for post-stroke cognitive outcomes.

## Introduction

Following stroke, some patients experience positive cognitive recovery outcomes, while others experience limited recovery or decline over time.^1–4^ This differential recovery has become a key research topic, as predicting individual recovery trajectories has potential to guide targeted interventions, conserve healthcare resources and improve care for those at risk of poor outcomes.^5–7^ While recent research has begun to identify clinical,^8,9^ cognitive^1,10–13^ and neuroimaging-based^14–16^ markers of recovery outcomes, additional work is needed to develop outcome prediction models which are both accurate and practical for real-world clinical settings.^16–18^

Recent work has indicated that lesion anatomy can be leveraged to predict chronic domain-specific outcomes.^19–21^ Reported predictive accuracy has varied depending on cohort characteristic (e.g. timing, age, severity and cognitive domains considered),^16,22,23^ but consensus is arising that lesion measures (particularly disconnection statistics) are associated with chronic outcomes. However, it is not yet clear whether these statistical relationships can be translated to support clinical prognostication.

Identifying lesion-behaviour correlates through lesion- and structural disconnection-mapping is a well-established approach offering valuable insight into causal neural correlates.^24,25^ This approach is conceptually different from, but occasionally conflated with, prediction-based analyses which explicitly measure the value of brain-behaviour relationships for predicting specific outcomes.^18,26^ Critically, the presence of brain-behaviour relationships alone does not necessarily imply that these relationships can be used to reliably distinguish between different outcomes.^18,26^ This means that specific analyses measuring predictive value through approaches such as out-of-sample validations and sensitivity analyses are needed to confirm the prognostic value of brain-behaviour relationships.^18,26^ Quantifying percent variance explained is a useful measure, but alone is not enough to infer predictive value as percent variance explained does not necessarily capture error magnitude, or interference from irrelevant variables (e.g. overfitting).^27^ Some studies have adopted out-of-sample prediction validation approaches^16,28^ which are the standard for evaluating predictive value, but this is not yet common practice. There are also many cases in which a predictive model explaining variance in chronic scores may not be making clinically meaningful predictions. For example, if very few patients exhibit impairment, models could achieve high accuracy by predicting high scores for all participants (regardless of their actual scores) but would not be providing meaningful predictions. Overall, there is a clear need for studies to more explicitly quantify the predictive value of brain-behaviour relationships.

Next, for a new predictive model to be useful in clinical environments, lesion metrics should provide additional information which improves diagnostic accuracy over and above other more easily available predictors. This is critical as lesion metrics take time and technical expertise to generate.^25,29,30^ To justify this additional workload, the information generated must provide a proportionate gain over predictors which can be more easily accessed. This issue is particularly relevant to predicting post-stroke cognitive outcomes, as there are many well-established clinical, demographic and behavioural outcome predictors.

For example, long-term cognitive impairment is associated with higher stroke age, lower socioeconomic status, incontinence and greater stroke severity.^31^ Factors such as pre-stroke cognitive decline, preexisting white matter disease, history of stroke and atrophy also predict cognitive outcomes.^8^ These metrics each interact with stroke specific factors such as lesion size, lesion location and stroke-specific cognitive impairment profile to modulate recovery outcomes.^8,11,16^ In this context, there is a clear need to differentiate between identifying factors which contribute to outcome predictions (e.g. through incremental explanatory power) and developing clinically useful prediction models which can inform clinical management.^9,32^ For example, an anatomical metric which significantly explains a small portion of variance in behaviour may be statistically significant and theoretically meaningful but may not be a strong enough to support accurate impairment predictions.^18^ Notably, a meaningful magnitude of predicted change is not established for cognitive outcomes.^33^ To evaluate whether lesion metrics are clinically useful prognostic indicators, it is important to determine whether these metrics provide additional predictive information over and above established predictors.

Importantly, post-stroke cognitive impairment is heterogeneous and complex.^34^ It is important for studies to capture, rather than mask, this heterogeneity of post-stroke cognitive impairment when modelling recovery trajectories. For example, stroke can selectively impact a range of dissociable cognitive domains, which are associated with differential recovery trajectories.^11,12,35–37^ It is not yet clear whether previously identified predictive relationships are consistent across a wider range of cognitive domains. The purpose of the present study was to identify neuroimaging predictors of persistent, domain specific cognitive impairment and to evaluate whether these measures provide additional prognostic utility relative to established risk factors. We aim to address this question using a multifaceted analytical approach by evaluating the strength of predictions supported by univariate brain-behaviour relationships (e.g. significant correlates identified in lesion mapping) and multivariate models incorporating whole-brain lesion status measures. Specifically, we aimed to determine whether lesion-based predictors can be used to effectively predict which patients will experience persistent cognitive impairment on each of the Oxford Cognitive Screen’s (OCS) subtask scores. This large-scale investigation provides a uniquely comprehensive assessment of the correlates of both acute and persistent cognitive impairment and provides a novel comparative evaluation of the utility of these imaging-based correlates to predict patient outcomes.

## Materials and Methods

### Patients

This is a retrospective analysis including 408 stroke patients recruited in the Oxford Cognitive Screening Programme (REC references:14/LO/0648, 18/SC/0550, 12/WM/00335), who completed the OCS at acute hospitalisation (< 30 days post-stroke) and chronic follow-up (∼6 months post-stroke). Participants were recruited consecutively during acute hospitalisation following a clinically confirmed stroke. Patients were excluded if they were unable to remain alert for the duration of a cognitive assessment (15 minutes) or if they were unable to provide consent.^38,39^ Participants were included in this analysis if they completed cognitive assessment at both time points and had available neuroimaging data of sufficient quality for lesion mapping.

Overall, 1098 patients completed acute cognitive assessment. Of these patients 436 did not complete the 6-month follow-up cognitive assessment. Additionally, 244 were excluded based on the neuro-imaging criteria: (i) no imaging within the relevant period (31 days of stroke), (ii) poor quality neuroimaging (e.g. motion artefacts, incomplete scans), (iii) evidence of multiple, temporally distinct strokes and (iv) no visible lesion damage on scan. Clinical and demographic information is reported in Table 1.

**Table 1:** Clinical and Demographic information. L = left, R = Right, F= Female

|  | **Sample Mean (SD, range)** |
| --- | --- |
| **Age** | 73.7 (12.4, 18-95) |
| **Sex** | 44.5% F |
| **Hand** | 7.97% L |
| **Years Education** | 12.2 (3.48, 7-28) |
| **Scan Type** |  |
| CT | 352 |
| MR | 56 |
| **Days to Assessment** | 4.3 (4.2, 0-31) |
| **Stroke Type** |  |
| Ischemic | 334 |
| Haemorrhage | 74 |
| **NIHSS** | 7.2 (6.2, 0-30) |
| **Barthel** | 13.2 (6.1, 0-20) |
| **Stroke Side** |  |
| Left | 164 |
| Right | 203 |
| Bilateral | 41 |
| **Stroke Volume** | 36.1 cm<sup>3</sup> (49.9, 0.007-303.7) |

### Behavioural Data

Cognitive deficits were identified using the OCS, a standard stroke-specific clinical screen assessing executive function, language, memory, number processing, visuospatial attention and praxis.^38,40^ OCS subtest scores were divided by the number of possible points to provide a proportion correct score which was comparable across all subtests. Persistent impairment was defined as cases in which patients were impaired on a subtest at acute and follow-up timepoints. Proportional behavioural scores for patients who do not exhibit significant impairment at both acute and chronic timepoints were constrained to 1 (ceiling) in analyses aiming to predict persistent impairment. This approach was taken to avoid conflating the expected, normal degree of variability in scores corresponding to intact behavioural performances with the changes in deficit severity observed in patients with abnormal cognitive scores.^40,41^ This approach is particularly critical here, given the restricted ranges of behavioural scores yielded by the OCS.^40^

### Neuroimaging

Lesions were manually delineated from routinely collected post-stroke neuroimaging (352 CT, 56 T2) by trained experts in line with a standard protocol.^42^ Lesion masks were smoothed at 5 mm full-width at half maximum in the z-direction, binarized (0.5 threshold), reoriented, warped into 1×1×1 mm stereotaxic space using Statistical Parametric Mapping and Clinical Toolbox functions^43,44^ and were visually inspected for quality. Anatomical statistics were generated via comparison to the Harvard-Oxford Cortical Atlas, Johns Hopkins White Matter Atlas, and, for disconnection analyses, the Schaefer-Yeo Atlas Parcellation (100 parcels, 7 Networks). In disconnection analyses, 35 subcortical and cerebellar parcels derived from the AAL and Harvard-Oxford Subcortical Atlas respectively were also included. These atlases were selected as they offer an optimum balance between parcellation complexity and anatomical interpretability.

### General Brain-Based and Behavioural Predictive Metrics

General brain-based metrics which are agnostic to detailed lesion anatomy were also considered. Cortical atrophy was rated using the Global Cortical Atrophy (GCA) scale.^45,46^ This scale assessed the severity of cortical atrophy in 13 brain regions by assigning each region an atrophy score of 0 (none), 1 (mild), 2 (moderate), or 3 (severe). Where regions were obscured by stroke lesions, regions were assigned the score of the homologous region within the undamaged hemisphere.^15^ Total GCA score was calculated by adding all region scores (GCA range = 0 – 39).

### Lesion Mapping Analyses

This study employs two distinct classes of analysis aiming to quantify brain-behaviour associations and to explore the value of derived metrics for predicting long-term cognitive outcomes. In terms of analyses establishing brain-behaviour relationships, ROI-level, network-level and multivariate voxel-level lesion mapping analyses were performed to identify of acute and persistent cognitive impairment. ROI- and network-level lesion mapping are mass univariate approaches in which regression analyses are conducted to determine whether the degree of damage to each defined anatomical parcel is significantly associated with impairment severity. Importantly, ROI-level lesion mapping evaluates both cortical and white-matter ROIs to provide a measure of both cortical and white matter damage (i.e. disconnectivity). In ROI-level lesion mapping, ROI damage scores were quantified by calculating the proportion of each atlas-defined ROI impacted by each lesion. In network-level lesion mapping, parcel-wise disconnection severities are estimated by determining the number of HCP-842 streamlines which bilaterally terminate in each pair of included grey matter parcels and calculating the proportion of these connections which are disrupted by each lesion.^29^

All analyses control for lesion volume (considered as a covariate of no interest in ROI and network analyses) and only voxels impacted in at least 3 patients were included. Where relevant, Bonferroni corrections for multiple comparisons were applied. Where no results survive this conservative correction threshold, less conservative 5% FDR corrections are used. This approach is employed because Bonferroni corrections can yield erroneously conservative lesion mapping results but are effective for identifying “core” correlates, while FDR corrections are less conservative, but have an elevated false positive risk.^47^ This flexible correction approach was selected to ensure the accuracy of generated prediction models was not constrained by strict corrections which may be appropriate in some cases (e.g. where large samples of impaired and spared patients are available), but not others (e.g. deficit categories with fewer impaired patients). Supplementary analyses were conducted to ensure this analytical choice did not significantly impact the prediction accuracy related conclusions reported in this study (Supplementary Table 3). All cases in which FDR corrections were applied are reported.

### Prediction Analyses

In terms of predictive value analyses, regression, receiver operating curve (ROC) and support vector regression (SVR) analyses were used to measure prognostic value. This study employed both univariate and multivariate analyses to explore the extent to which chronic cognitive status can be predicted by lesion, brain health and behavioural scores. First, univariate analyses were conducted to evaluate whether significant correlates identified in lesion mapping analyses are able effectively identify patients with persistent cognitive impairments. In univariate analyses, patient lesion masks were overlayed with significant correlates of persistent impairment to generate an “impact score” representing the extent to which critical correlates were damaged in each patient (0 = no overlap between lesion and critical correlates, 1 = complete lesion coverage of critical correlates). In ROI-level predictive models impact scores were the proportion of lesioned critical ROI voxels by the total number of critical ROI voxels. For network-level analyses, patients’ average disconnection values within critical connections were used as the impact score.

First, regression analyses were conducted to assess whether all considered predictors (ROI impact score, network impact score, acute behaviour, GCA score) were significantly associated with chronic cognitive status when considered independently. Bayesian regression analyses were also applied to provide an explicit measure of the strength of evidence in support of associations between predictors and chronic outcomes. These analyses employed alternative hypotheses with JZS prior (scale factor = 0.707) and a null hypothesis prior set at chance level.

Next, Receiver Operating Curve (ROC) analyses were conducted to assess the diagnostic performance of all considered predictors. Each predictor value was scaled between 0-1, and a range of impairment threshold cut-offs were applied (100 values evenly spaced between 0 and 1). The relationship between sensitivity and specificity for each of these cut-offs was plotted and area under curve (AUC) values were calculated. For diagnostic tools, tests with AUC values of 0.9-1 are considered excellent, 0.8-0.89 are good, 0.7-0.79 are fair, 0.51-0.69 are poor and AUC of <0.5 are considered to be of no diagnostic value.^48^

Next, multivariate prediction analyses were conducted to quantify prediction accuracies when predictors were considered concurrently. Specifically, support vector regression analyses were conducted to identify and evaluate the strength of multivariate predictive relationships between lesion anatomy and cognitive outcomes. Support vector regression is a robust machine learning approach which predicts continuous outcome variables (e.g. cognitive scores) by building multivariate functions based on input variables.^49,50^ These multivariate analyses do not employ lesion impact scores considered in univariate analyses but instead consider whole-brain profiles of ROI/network-level damage. The predictive accuracy of five distinct SVR models was evaluated. Models 1-4 were trained to predict cognitive outcomes based on ROI impact statistics, network-level disconnection, acute OCS performance and GCA region-level scores. Model 5 considered all these predictors together.

Each support vector regression model was trained to predict cognitive outcomes based on 75% of data. Model accuracy (root mean square error (RMSE)) was calculated within the remaining left-out 25% of data (linear kernel, epsilon regression). Models were tuned to select optimal hyperparameters (cost and gamma) prior to training. This process was repeated 100 times for each model, with overall model accuracy being average RMSE across iterations (lower RMSE = more accurate predictions). For each cognitive outcome, differences in accuracy between behavioural, ROI, network-level, brain health and combined values were identified using ANOVA analyses and post-hoc Tukey HSD tests.

## Results

### Behavioural Results

Acute and follow-up OCS performance descriptives are reported in Table 2.

**Table 2:**
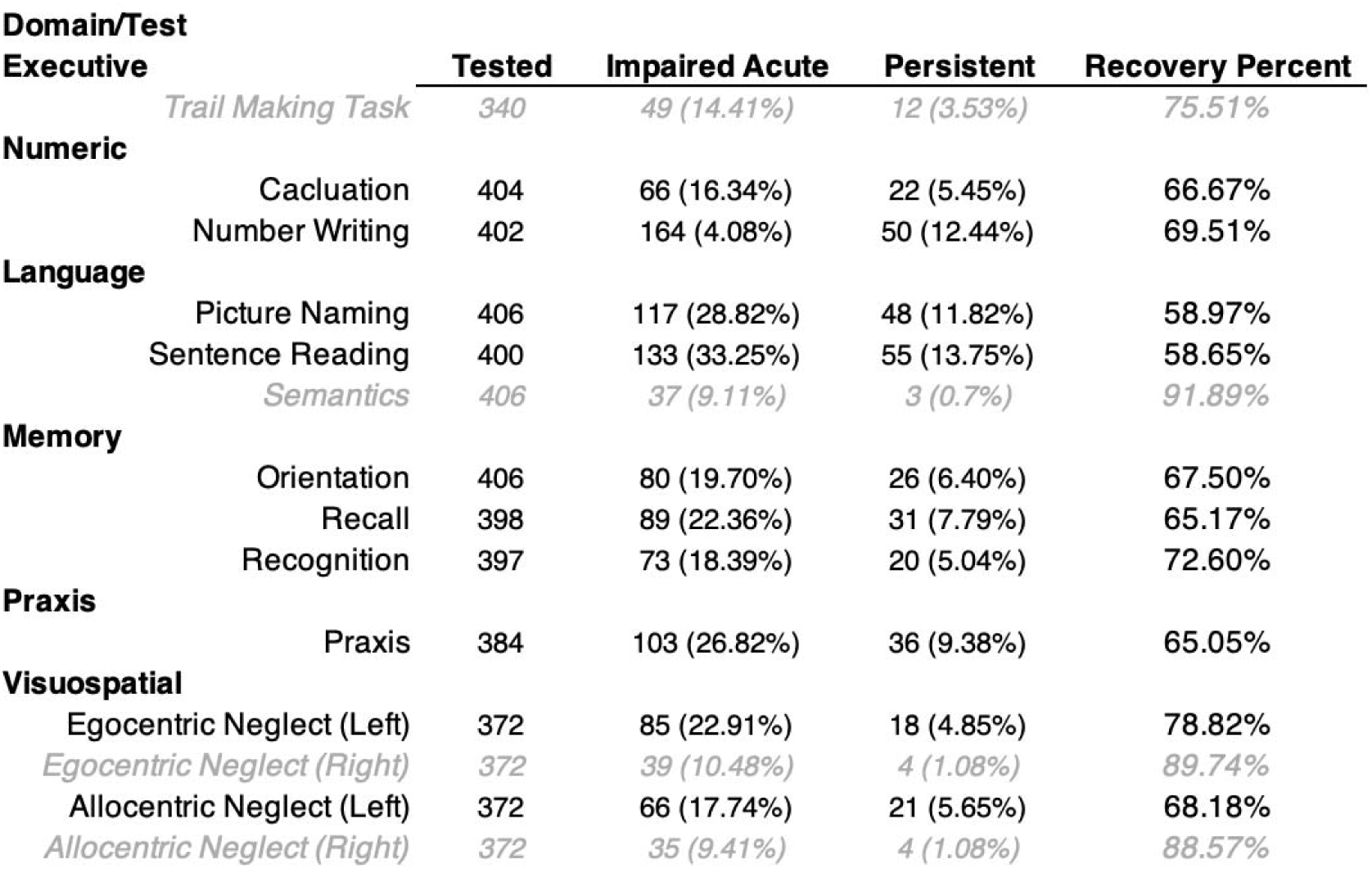
Behavioural testing results. Tested reports the number of patients tested at both acute and follow-up time points. This sample’s cognitive impairment prevalence and recovery rate is in line with continuously recruited stroke cohorts reported in previous work.^11^ Results for subtests with <15 persistently impaired participants (indicated in grey italics) are reported in supplementary materials.

| Domain/Test | Tested | Impaired Acute | Persistent | Recovery Percent |
| --- | --- | --- | --- | --- |
| **Executive** |  |  |  |  |
| *Trail Making Task* | *340* | *49 (14.41%)* | *12 (3.53%)* | *75.51%* |
| **Numeric** |  |  |  |  |
| Cacluation | 404 | 66 (16.34%) | 22 (5.45%) | 66.67% |
| Number Writing | 402 | 164 (4.08%) | 50 (12.44%) | 69.51% |
| **Language** |  |  |  |  |
| Picture Naming | 406 | 117 (28.82%) | 48 (11.82%) | 58.97% |
| Sentence Reading | 400 | 133 (33.25%) | 55 (13.75%) | 58.65% |
| *Semantics* | *406* | *37 (9.11%)* | *3 (0.7%)* | *91.89%* |
| **Memory** |  |  |  |  |
| Orientation | 406 | 80 (19.70%) | 26 (6.40%) | 67.50% |
| Recall | 398 | 89 (22.36%) | 31 (7.79%) | 65.17% |
| Recognition | 397 | 73 (18.39%) | 20 (5.04%) | 72.60% |
| **Praxis** |  |  |  |  |
| Praxis | 384 | 103 (26.82%) | 36 (9.38%) | 65.05% |
| **Visuospatial** |  |  |  |  |
| Egocentric Neglect (Left) | 372 | 85 (22.91%) | 18 (4.85%) | 78.82% |
| *Egocentric Neglect (Right)* | *372* | *39 (10.48%)* | *4 (1.08%)* | *89.74%* |
| Allocentric Neglect (Left) | 372 | 66 (17.74%) | 21 (5.65%) | 68.18% |
| *Allocentric Neglect (Right)* | *372* | *35 (9.41%)* | *4 (1.08%)* | *88.57%* |

### ROI-level lesion mapping

ROI-level lesion mapping identified regions significantly associated with acute impairment in 9/10 OCS subtests (all *P* < 0.0007) (Figure 1). Significant correlates of persistent impairment were identified within 7/10 OCS subtests (all *P* < 0.0004). There was overlap within the correlates predicting acute and persistent impairment within sentence reading, picture naming, number writing and left egocentric/allocentric neglect. Within all other considered subtests, acute and persistent impairments were linked to damage to distinct, non-overlapping regions. Full anatomical descriptive statistics for all significant ROIs are available in Supplementary Table 2. All correlates survived Bonferroni corrections, except for the correlates of acute orientation (FDR corrected).

**Figure 1:**
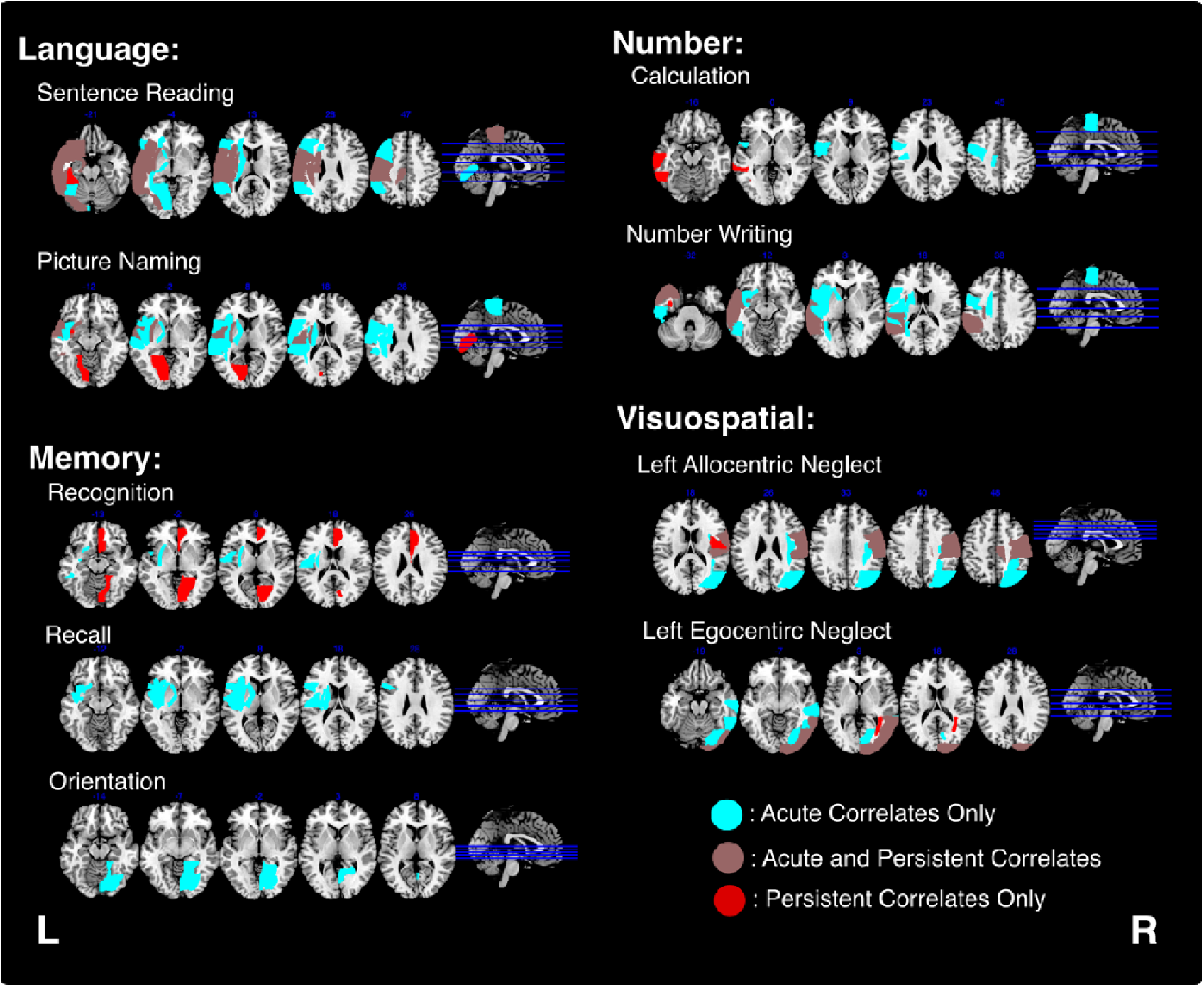
ROI-level correlates of acute and persistent impairment. Correlates predicting acute, but not persistent impairment are blue. Regions predicting both acute and persistent impairment are brown and areas predicting persistent, but not acute impairment are red. The location of MNI axial slices are denoted by the blue lines on the sagittal slice.

### Network-Level Lesion Mapping Results

Network-level lesion mapping identified disconnectivity patterns associated with acute impairment on all OCS subtests (all *P* < 0.0004) and with persistent impairment for 9/10 OCS tests (all *P* < 0.002). Network-level results for acute orientation, praxis and calculation subtest scores survived FDR corrections, but not Bonferroni corrections. Similarly, results for persistent verbal recall, praxis, number writing, calculation and left egocentric neglect impairment was significant when FDR corrections (but not Bonferroni corrections) were applied. Acute and persistent network-level correlates were found to overlap within OCS language tasks, verbal recall, number writing, calculation and left allocentric neglect.

### Lesion Location Information Predicts Persistent Cognitive Impairment

Regression analyses (n = 10) were conducted to investigate whether each considered predictor type (ROI impact score, network impact score, acute behaviour, GCA score) was significantly associated with chronic cognitive impairment when considered independently. ROI and network impact scores were both significantly associated with chronic cognition in all cases where significant correlates of persistent impairment were identified, except for network impact scores in the gesture imitation subtests. ROI impact scores explained between 3% (Calculation) and 18% (Sentence Reading) variance on OCS subtests (mean = 8%, SD= 5%), with all Bayes Factors indicating strong evidence in favour of an association. Similarly, network impact scores explained between 1% (Recognition) and 13% (Sentence Reading) of variance in chronic scores (mean = 6%, SD = 4%). Bayes Factors indicated strong evidence of an association for 7/9 subtests, weak evidence in one subtest (Recognition) and moderate evidence for the absence of an association in one subtest (Praxis).

Conversely, GCA total score was significantly associated with chronic cognition in 7/10 subtests, explaining between 1% (Calculation, Number Writing) and 7% (Recognition) of variance. Bayes factors indicated moderate evidence against an association in two tasks, and weak, moderate and strong evidence in two, one and five tasks respectively. Acute behaviour exhibited the strongest associations with chronic cognition, explaining between 9% (Gesture Imitation) and 29% (Picture Naming) of variance (all *P* < 0.001). Bayes factors indicated strong evidence of an association in all cases.

### Lesion anatomy data has reduced prognostic utility compared to acute behavioural data

Next, ROC analyses were conducted to evaluate whether considered predictors can identify patients with persistent impairment. All ROI and network impact scores showed low diagnostic accuracy under conventional AUC thresholds (max AUC = 0.657) (Figure 3, Table 4). Across all subtests the best predictor of persistent impairment was acute behaviour for 9/10 OCS subtests, and GCA for the remaining 1/10 subtest.

**Figure 2:**
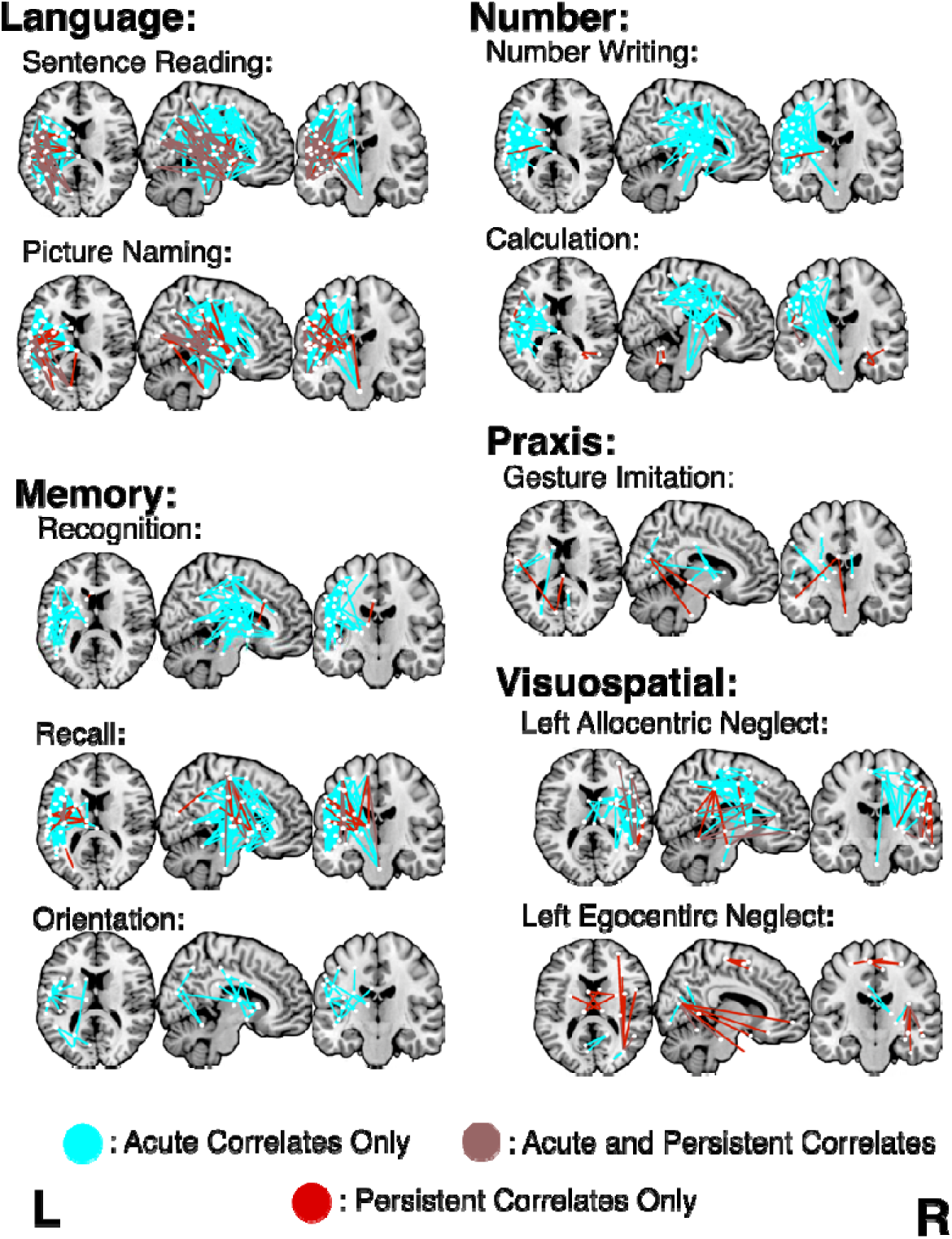
Network-level correlates of acute and persistent impairments. Network-level correlates of acute and persistent cognitive impairment as reported by OCS subtests. Network edges associated with only acute behavioural scores are shown in blue, edges associated with both acute and persistent impairment are shown in brown and edges associated with persistent, but not acute impairment are shown in red. Axial slices are MNI z = 14, coronal are MNI y = -16, and sagittal are MNI x = 8. Names, MNI coordinates, and lesion mapping statistics for each node are openly available (https://osf.io/9zkyh/).

**Figure 3:**
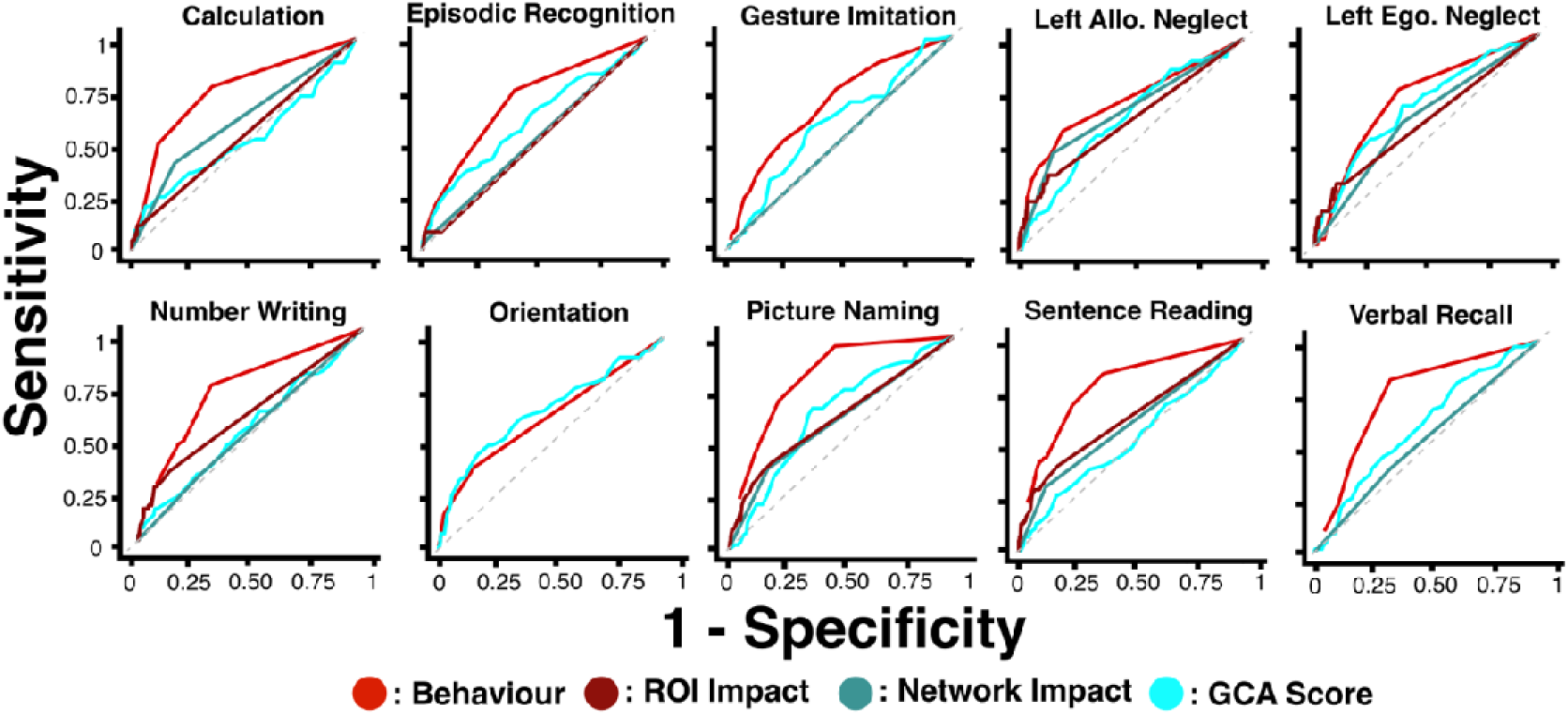
Diagnostic accuracy of prediction models. ROC curves illustrating the diagnostic accuracy of each considered predictor across all OCS tests. Line colour represents predictor type. The best performing predictive metric is listed in the bottom right corner of each plot.

**Table 3:** Multivariate regression results. Results statistics for multivariate regressions aiming to determine whether lesion location scores add value when considered alongside general brain-based predictors.

|  | Acute Behaviour |  | ROI Correlates |  | Discon. Correlates |  | Atrophy |  |
| --- | --- | --- | --- | --- | --- | --- | --- | --- |
|  | R2 | BF | R2 | BF | R2 | BF | R2 | BF |
| **Language** |  |  |  |  |  |  |  |  |
| Picture Naming | 0.29 | $3.27 \times 10^{28}$ | 0.07 | $5.09 \times 10^5$ | 0.07 | $4.98 \times 10^5$ | 0.04 | $1.28 \times 10^3$ |
| Sentence Reading | 0.21 | $4.7 \times 10^{18}$ | 0.18 | $7.41 \times 10^{15}$ | 0.13 | $2.79 \times 10^{11}$ | 0.00 | 0.14 |
| **Memory** |  |  |  |  |  |  |  |  |
| Orientation | 0.09 | $4.49 \times 10^7$ | | | | | 0.05 | $2.29 \times 10^3$ |
| Recognition | 0.12 | $1.26 \times 10^9$ | 0.02 | 10.76 | 0.01 | 2.00 | 0.07 | $9.87 \times 10^4$ |
| Recall | 0.16 | $2.71 \times 10^{13}$ | | | 0.02 | 10.69 | 0.02 | 15.81 |
| **Number** |  |  |  |  |  |  |  |  |
| Calculation | 0.16 | $6.73 \times 10^{13}$ | 0.03 | 93.23 | 0.07 | $1.29 \times 10^5$ | 0.01 | 1.50 |
| Number Writing | | | 0.10 | $2.02 \times 10^8$ | 0.04 | 252.51 | 0.01 | 1.67 |
| **Visuospatial** |  |  |  |  |  |  |  |  |
| Left Egocentric Neglect | 0.11 | $3.28 \times 10^8$ | 0.09 | $2.01 \times 10^7$ | 0.08 | $8.54 \times 10^5$ | 0.02 | 6.77 |
| Left Allocentric Neglect | 0.18 | $7.81 \times 10^{14}$ | 0.09 | $2.21 \times 10^7$ | 0.10 | $1.31 \times 10^8$ | 0.00 | 0.38 |
| **Praxis** |  |  |  |  |  |  |  |  |
| Gesture Imitation | 0.09 | $2.91 \times 10^6$ | | | 0.00 | 0.32 | 0.05 | $2.58 \times 10^3$ |

**Table 4:**
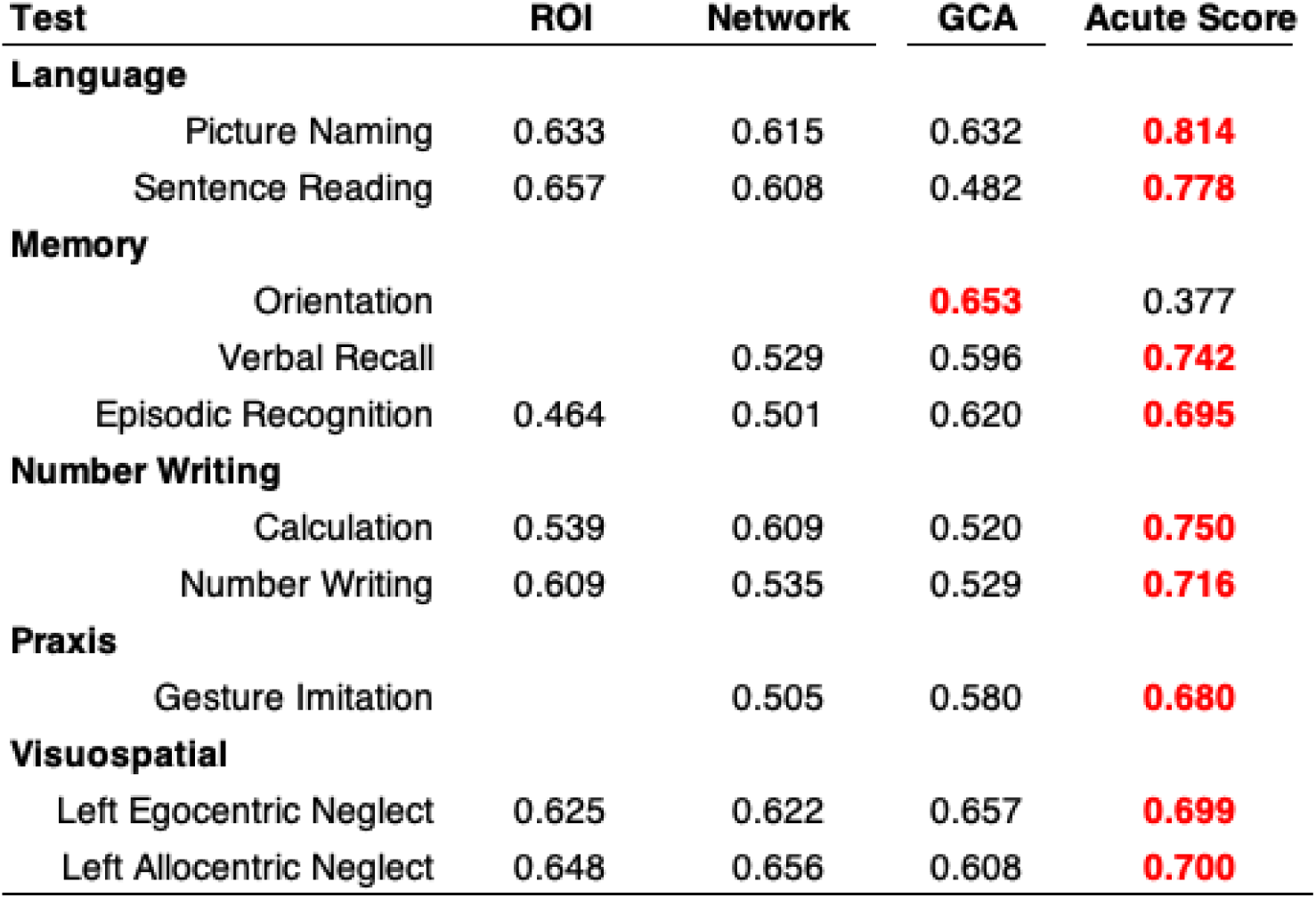
AUC values for predictive models. AUC values for each considered predictor across all OCS subtests. The highest AUC value for each subtest is highlighted in red.

Finally, SVR analyses were conducted to identify potential multivariate predictive relationships between lesion anatomy and cognitive outcomes (Figure 4). Across all considered tests, prediction accuracy was highest for SVR models trained on behaviour alone (mean RMSE = 0.018), followed by models trained using region-specific GCA scores (mean RMSE = 0.041), ROI impact data (mean RMSE = 0.102), models considering all data (mean RMSE = 0.176) and network-level disconnection data (mean RMSE = 0.180). ANOVA analyses indicated the presence of significant differences between different model types in all cases (all *P*-values < 0.001).

**Figure 4.**
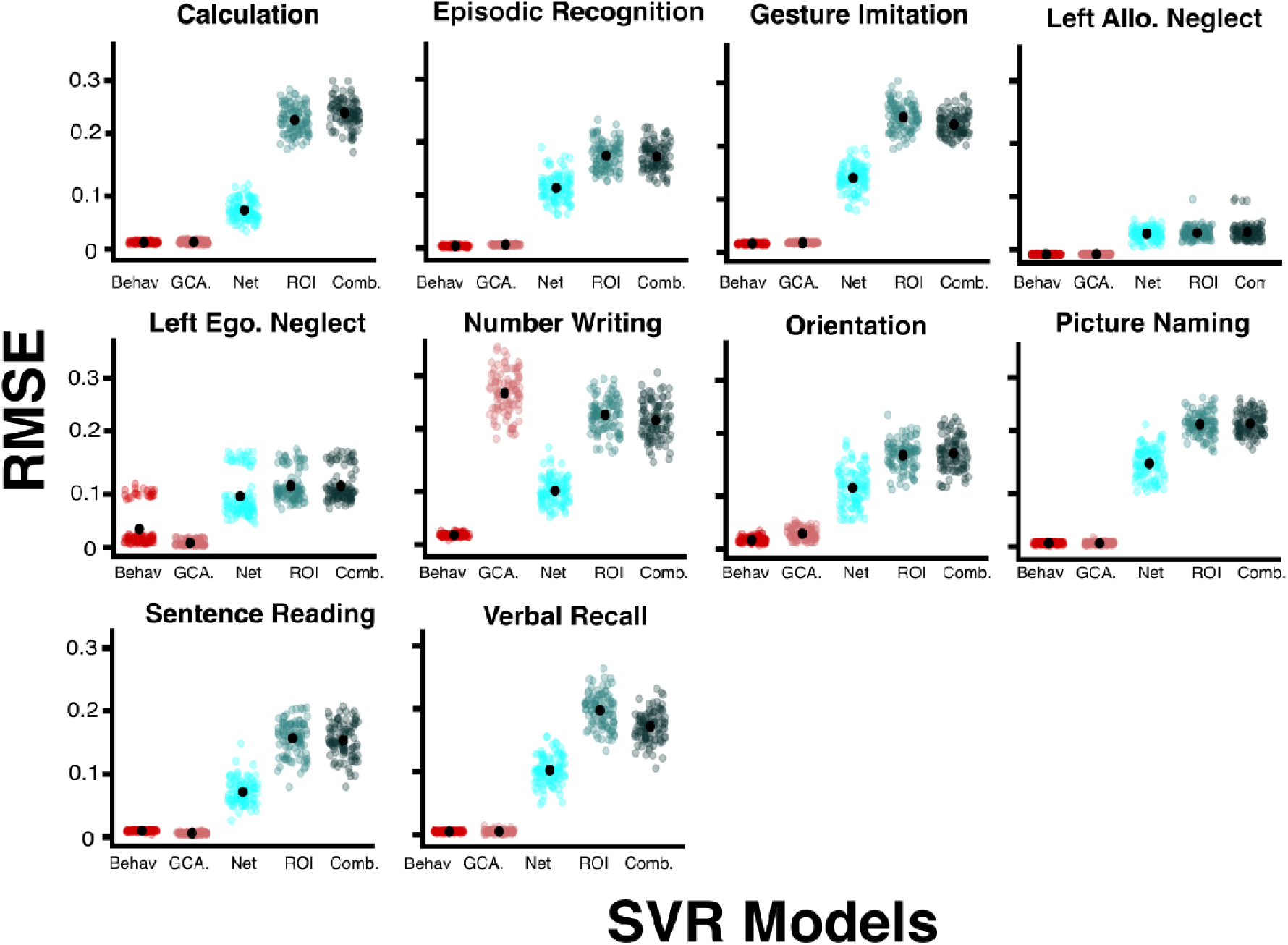
Prediction accuracy for SVR models: Prediction accuracy for SVR models trained to predict chronic OCS scores based on acute behaviour (Behav.), region-level GCA scores (GCA), ROI impact scores (ROI), network-level disconnection probabilities (Net.) and all these data sources combined (Comb.). Accuracy is reported in terms of root mean squared error, with lower RMSE representing more accurate predictions.

In all cases, SVR models trained to predict outcomes based on behavioural data and region-level GCA scores both significantly outperformed models trained on ROI data, network-level disconnection and models trained using all data types combined (all *P* < 0.001). However, models trained on acute behaviour were only significantly more accurate than models trained on region-level GCA data in two cases (Orientation, Number Writing), performed significantly worse than GCA scores in predicting chronic left egocentric neglect, and did not perform significantly differently from GCA models in the remaining 7 OCS tests. In terms of lesion-based models, ROI data SVR models outperformed both network-level and combined data models in all cases, except for left allocentric neglect scores, where these models were not significantly different in terms of accuracy. Network-level model accuracy significantly outperformed combined model performance in predicting chronic calculation impairment (*P* < 0.001). Conversely, combined data models significantly outperformed network-level models in two cases: Praxis and Verbal Recall (*P* < 0.001)

ROI based SVR models significantly outperformed both network-level and combined data SVR models in all cases (*P* <0.001), except left allocentric neglect, where no differences were present between ROI models and network-level (*P* = 0.837) or combined models (*P* = 0.198). Models trained on combined data outperformed network disconnection-based models in two OCS subtests (Gesture Imitation and Verbal Recall, *P* < 0.001) but performed significantly worse than network disconnection models within the calculation subtest (*P* < 0.001). Overall, these results are in line with univariate lesion mapping analyses in suggesting that both lesion data and behavioural data can be leveraged to predict chronic outcomes, but acute behavioural data is the strongest predictor of chronic cognitive status.

## Discussion

This study provides a comprehensive evaluation of the incremental value of lesion location metrics relative to general brain-based predictors and acute behaviour for predicting post-stroke cognitive outcomes. These results provide novel insight into the detailed neuroanatomy of both acute and persistent cognitive impairment across a diverse range of commonly impacted domains. In line with previous studies, statistically significant lesion and disconnection correlates associated with long-term cognitive outcomes were identified. However, this anatomical detail was found to have limited prognostic benefit when considered alongside simple brain-health metrics and acute behaviour as implemented here using routine clinical imaging and standard mapping pipelines. Acute behaviour was the single strongest predictor of chronic cognitive status in both univariate ROC analyses and in multivariate SVR analysis. Total GCA score was not a useful prognostic indicator in ROC analyses, but region-level GCA scores were able to provide accurate predictions of chronic cognitive outcomes in SVR analysis. Overall, the results of this study suggest that the prognostic utility of complex anatomical metrics (as quantified here) has limited incremental value compared to more readily available prognostic indicators.

### Identified correlates of domain-specific impairments align with past work

The identified correlates of acute impairments largely agree with those described in past work.^51,52^ In line with previous work, language-domain tasks (picture naming, sentence reading) were associated with damage to and disconnection within left fronto-temporal regions. ^53–56^ Numerical cognition deficits (number writing, calculation) were also associated with left hemisphere damage and disconnection.^57,58^ ROI-level correlates of left neglect were centred in right parietal regions, and these deficits were associated with widespread patterns of right-hemisphere disconnection.^59–61^ However, in contrast to previous literature, acute orientation impairment was significantly associated with damage to the right lingual gyrus and occipital fusiform cortex in ROI-level analyses. Orientation impairment has generally been linked to left hemisphere correlates in either MCA territory^62^ or more posterior regions such as the occipitotemporal and precuneal cortex.^63^ Notably, this study’s ROI-level results for acute orientation survived FDR but not Bonferroni corrections for multiple comparisons, indicating that the identified relationship between right posterior damage and orientation is relatively weak.^30,64^

The correlates of persistent domain-specific cognitive impairments are not well established. Some studies have reported analyses comparing correlates of acute and chronic left neglect^65,66^ and post-stroke language impairment^67,68^, but similar analyses have not been conducted for the majority of considered cognitive tasks. This study’s network- and ROI-level analyses found that acute and persistent impairment on individual cognitive tasks are often associated with different patterns of lesion damage. Some degree of correlate variability is expected due to the expected changes in behavioural scores over time^30^. However, the acute and persistent impairment were sometimes associated with strikingly different lesion patterns including correlates which reversed lateralisation (e.g. episodic recognition). This finding emphasises the need for future lesion mapping analyses to avoid combining acute and chronic behavioural scores, as this can introduce potentially confounding anatomical variability.^25,30^ Future, more detailed research, is needed to explore the mechanisms which may be driving differences in lesion anatomy over time.

### Why do prognostic estimates differ across lesion-based prediction studies?

Recent literature has reported that disconnection profile can be leveraged to predict long-term cognitive outcomes.^14,16,20,22,23^ While this study’s results may seem to conflict with this past work, several important nuances should be considered. First, past studies have identified statistically significant relationships between specific patterns of damage and disconnection and performance on neuropsychological measures.^14,20,21^ Importantly, these results are replicated in the present study which found significant relationships between lesion anatomy or disconnection and chronic performance in 9/10 considered OCS measures. The strength of these relationships is indeed lower in this study relative to previous work, but this difference is likely indicative of difference in cohort demographics and detail of the employed neuropsychological tests. This study also supports the previously proposed conclusion that disconnection profile is associated with chronic cognition, as damage to white matter tracts was significantly associated with persistent cognitive impairment in many OCS subtests within ROI-level analyses.^14,16,23^ However, this study differs from this past work in its aim to explicitly evaluate whether these identified statistically significant relationships are of sufficient strength to act as useful prognostic indicators in clinical settings.

Despite the identified significant relationships between lesion anatomy and chronic behaviour, the strength and prognostic utility of these metrics was low compared to acute OCS and region-specific global cortical atrophy scores. Acute cognition has been consistently found to be a strong predictor of chronic cognitive status^8,11,37,69,70^ and recent work on the OCS has shown that acute behaviour based individual level prediction modelling may be viable^10^. Past research has also identified that atrophy severity is a strong predictor of poor, non-cognitive stroke outcomes such as functional status,^71^ unfavourable anticoagulation responses^72^ and higher levels of disability.^73^ In terms of cognition, atrophy has been associated with worse NIHSS scores and executive functioning 6-weeks post stroke.^73^ Estimated brain age derived from MRI scans, which captures atrophy, has recently emerged as a promising biomarker of long-term stroke outcomes.^74,75^ Brain age predicts aphasia recovery outcomes, but the value of this predictor has not yet been extensively evaluated within other cognitive domains.^74^ A previous study reporting on a subset of the cohort reported here found that visual ratings of global cortical atrophy were strongly associated memory and executive function abilities 6-months post-stroke.^15^ Overall, the results of this study build upon this past work by highlighting atrophy severity as a promising prognostic indicator for post-stroke cognitive outcomes. This result is particularly encouraging as visual ratings of atrophy (such as GCA region scores) can be quickly generated in clinical settings without the use of specialist software.

In terms of acute behaviour, previous imaging-derived predictive models have generally not incorporated detailed baseline behavioural performance or have controlled for this by including domain-general scores summarising impairment severity (or demographics) rather than including domain-specific cognitive scores. In cases where more detailed baseline measures were considered, early behavioural scores have been found to explain larger portions of variance in chronic scores compared to imaging metrics.^20,21^. Sperber et al.^18^ illustrated that statistical relationships between lesions and outcomes do not necessarily translate into useful prognostic indicators, especially in cases where model fit has not been evaluated on external cohorts. Many previous studies which have reported that lesion metrics provide strong predictions of chronic cognitive status have not explicitly evaluated whether these results can be replicated in external cohorts. Notably, Talozzi et al.^16^ developed a disconnection-based predictive model which was able to provide above change (R^2^ = 0.18-0.20) predictions of long-term impairments in two external samples. However, subsequent studies have indicated that the predictive power of this approach may decrease in cases where cognitive outcomes are measured at different timepoints (e.g. 6-months and 5+ years) post-stroke.^22^

This result highlights a key issue which may help explain why predictive accuracy was low in the present study. Mainly, this study considers a diverse and representative cohort including older patients with large and spatially diverse lesions, comorbid atrophy as well as higher levels of acute impairment (mean acute NIHSS = 7.2). These factors have been demonstrated to modulate the specific profile of cognitive impairments expressed after stroke, independently of lesion anatomy.^76^ This inclusive approach differs from previous studies which have included selective, ‘clean’ cohorts with younger average ages (e.g. mean age = 50-65) and less severe strokes as indicated by lesion volumes (e.g. mean volume = 3-7 cm^3^).^23^ Past work has indicated that brain-behaviour relationships are strongest in patients with low levels of atrophy and good white matter integrity (i.e. younger stroke patients), suggesting that predictive models which are trained on younger or more selective stroke samples may not generalise to representative cohorts of stroke survivors. Disconnection metrics may also be sensitive to variability in lesion size, cohort heterogeneity and registration noise. These implications are critical to consider in the context of future predictive models, as models which are not trained and tested in representative cohorts may limited generalisability.

### The value of individual recovery predictors depends on what is being predicted

Notably, the diagnostic value of predictive metrics varied across cognitive domains. While ROI and disconnection measures yielded comparatively high AUCs when used to predict language and neglect deficits, these metrics preformed more poorly at predicting persistent impairment on memory domain tasks. These findings are broadly in line with past work demonstrating that variance in aphasia and neglect recovery outcomes can be explained by lesion anatomy.^20,21^ Conversely, past work has highlighted that much of the variance in post-stroke memory can be linked to general brain health measures and pre-morbid cognitive status than to the specific location of stroke lesions.^15,74,76^ In line with past work, GCA severity was most strongly associated with persistent memory domain impairments and was the single strongest predictor of orientation impairment in ROC analyses.

Acute behavioural scores were the single strongest predictor of persistent cognitive impairment, but the predictive value of this metric was not always high. In univariate analyses, acute behavioural scores explained only 9% of variance in chronic praxis and orientation impairment. This low explanatory power is likely linked to the comparatively low incidence of chronic impairment on these subtasks. Where prevalence of chronic impairment is low, models are underpowered and less stable.^3^ Notably, SVR analyses indicated that considering the lesion metrics considered here in addition to behavioural data did not improve the accuracy of predicted chronic cognitive outcome scores. The magnitude of difference between behaviour-only and combined models varied across OCS subtests, but models including more lesion predictors (e.g. network-level and combined models) were consistently outperformed by simpler models (behaviour and ROI-level models). Increasing the number of predictor variables increases the chance that models will be overfit to training datasets and therefore yield poor results when applied to held-out testing data.^77^ The risk of overfitting was minimised by optimising the model hyperparameters determining decision boundary sensitivity and model complexity, but this procedure does not eliminate all risk.^78^ Past research has found that adding more detailed lesion anatomy information (such as disconnection) does not always improve upon predictions generated by considering simpler anatomical descriptive statistics.^28^ The results of this study are in line with this previous implication.

### Incremental improvement alone isn’t good enough for real-world diagnostic predictions

Generating lesion anatomy statistics is an intensive process which requires high-level technical expertise, the use of specialist analytical software and significant time resources.^25,25,29,42^ Because of this, clinicians do not generally have access to the standard-space lesion mask data which would be needed to the type of anatomical impact scores used in this study. For lesion-location information to be useful to clinicians, the cost of generating these detailed metrics must be outweighed by the added prognostic information provided by this data. It is possible that some of this work will be automated in future, but then the needed computational power and processing time will still require justification. The findings of this study suggest that this criterion is not currently met. In this study, the predictive value of lesion-based measures (considered in isolation) was always outweighed by more accessible data sources including visual ratings of brain health and acute behaviour. This result is critical as statistical significance entails little practical utility if the considered prognostic metric cannot effectively distinguish between populations of interest.

Importantly, this result should not discourage future investigations into prognostic anatomical relationships but instead should encourage more thorough evaluations of whether identified predictive relationships provide additional explanatory power. Establishing predictive brain-behaviour relationships will continue to be highly relevant to theoretical discourse, but it is important that any potential practical utility of these relationships is thoroughly evaluated and interpreted with appropriate caution. Overall, future studies should aim to continue to develop accurate, detailed and generalisable prognostic models capturing stroke recovery trajectories. However, until these models can be demonstrated to provide substantial improvements, prognostic evaluations should focus on more readily available information sources.

### Limitations

Regardless of whether analyses are univariate or multivariate, lesion-mapping results are subject to some degree of results mislocalisation.^30,79–81^ This is because lesion locations are inherently non-random as their location is ultimately determined by the brain’s underlying vasculature structure ^80^. This study uses routinely available clinical imaging and behavioural data. While routinely collected stroke imaging has been repeatedly demonstrated to be of sufficient quality to localise correlates of post-stroke impairment,^51,82^ this imaging lacks the resolution necessary to map more fine-grained brain-behaviour relationships with potential prognostic value (e.g. specific branches of white matter tracts or neurotransmitter systems). Similarly, the OCS is a quick bedside cognitive screen which does not capture the same level of cognitive detail as more in-depth and lengthy neuropsychological assessment batteries.^40^ While this screen does broadly measure the most common cognitive consequences of stroke, it is possible that the recovery trajectories of other, more fine-grained deficits (e.g. subdivisions of aphasia) may be more clearly linked to lesion location. This study also uses a sample which is comparable to the stroke population in terms of age and impairment levels, meaning that it is possible that results may not generalise to younger or more selective (i.e. mild) stroke samples. Despite the large sample included here, the prevalence of some domain-specific cognitive impairments was low at the chronic timepoint (n with persistent impairment = 15 – 55). Where prevalence of chronic impairment is low, models are underpowered and less stable, meaning that additional work is needed to validate these findings in large samples of participants exhibiting specific types of chronic PSCI.^3^ Future research can aim to investigate these possibilities and further explore how lesion location information can be applied to predict post-stroke recovery trajectories.

## Data Availability

All data and code associated with this project is openly available on the Open Science Framework (https://osf.io/9zkyh/).

## Acknowledgements

This project was supported by the Oxford Cognitive Screening Programme. Specifically, we would like to acknowledge Dr. Georgina Hobden for her contributions to generating the cortical atrophy rating used in this study.

## Supplementary Materials

### Results from OCS Tests with <15 persistently impaired patients

**Supplementary Figure 1:**
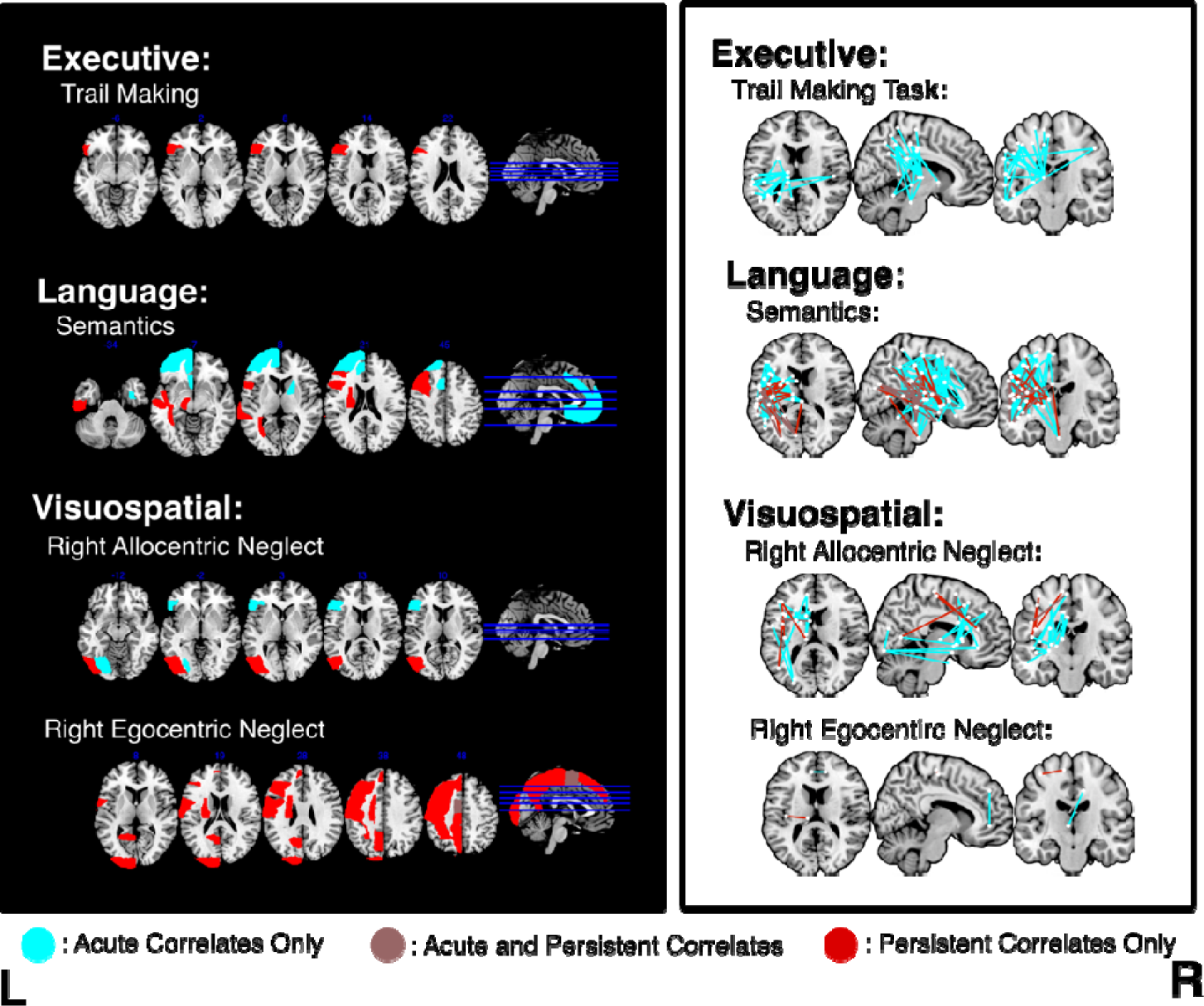
ROI-level and network-level lesion mapping results from OCS subtests with <15 persistently impaired patients. Within ROI results, the location of MNI axial slices are denoted by the blue lines on the sagittal slice. Edges/ROIs associated with only acute behavioural scores are shown in blue, edges associated with both acute and persistent impairment are shown in brown, and regions/edges associated with persistent, but not acute impairment are shown in red. Within network-level results, axial slices are MNI z = 14, coronal are MNI y = -16, and sagittal are MNI x = 8. Names, MNI coordinates, and lesion mapping statistics for each node are openly available (https://osf.io/9zkyh/).

### Multivariate Lesion Mapping Analyses and Results

Multivariate lesion mapping analyses were conducted using sparse canonical correlations (SCCAN). SCCAN builds a best-fit model by assigning weights to voxel distributions which capture multivariate associations between damage patterns and behavioural scores. The proportion of voxels retained in models is defined through a sparseness optimisation procedure ^83^. Resultant models are only considered valid if they generate statistically significant behavioural classifications within a four-fold cross validation approach. This method has been validated and has been shown to offer increased results accuracy compared to traditional mass-univariate lesion mapping approaches ^78^.

SCCAN lesion mapping analyses identified significant correlates of acute impairment within 8/15 considered OCS subtests (Supplementary Figure 2). No significant correlates of persistent impairment were found. Full SCCAN statistics including number of significant voxels, voxel weight ranges, and sparseness values are reported in Supplementary Figure 1. As no significant correlates of persistent impairment were identified in SCCAN analyses, these results were not used in prediction analyses.

**Supplementary Figure 2:**
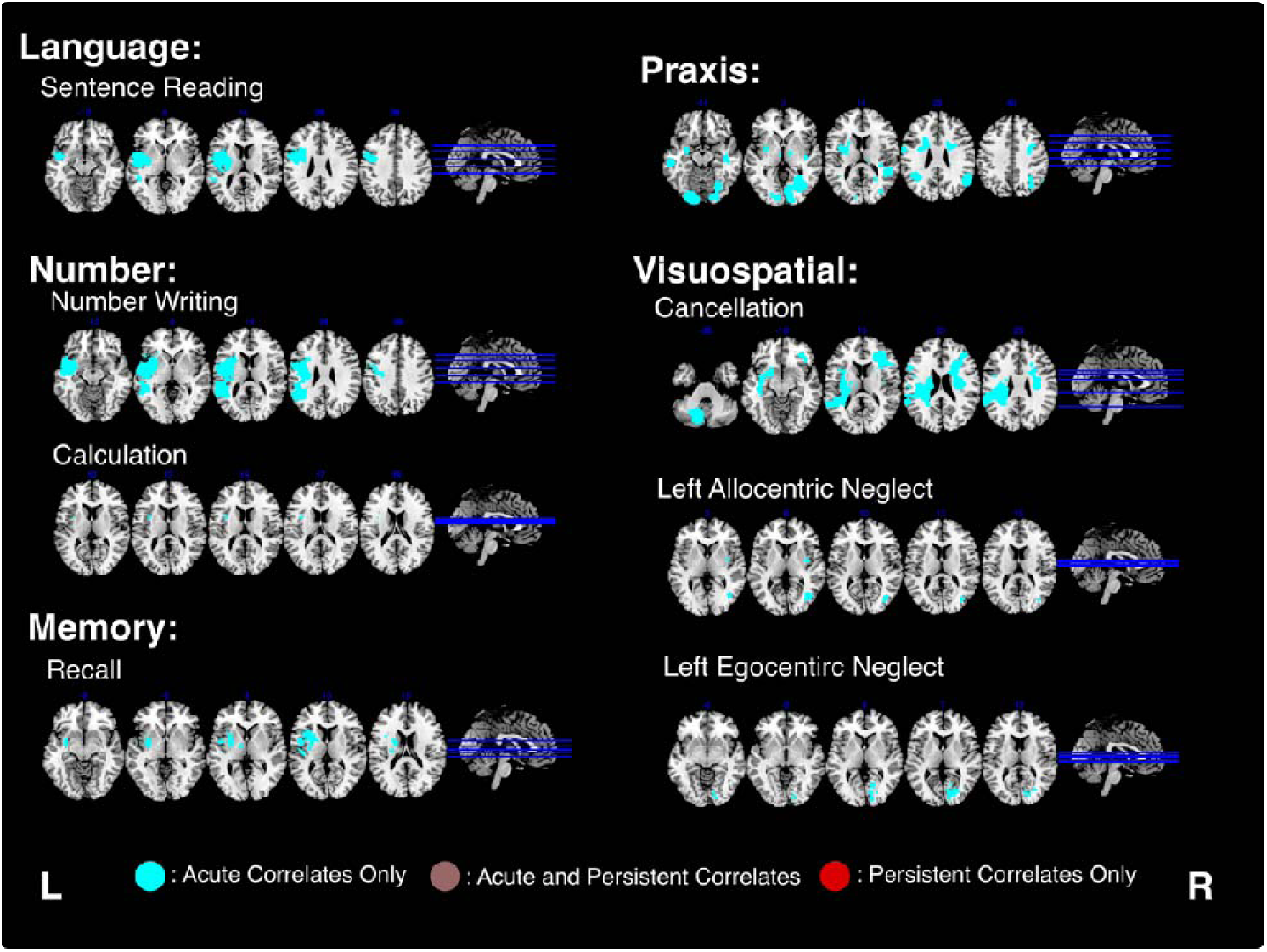
Results from SCCAN lesion mapping analyses. Voxels associated with only acute impairment are shown in blue. No correlates of persistent impairment were identified in SCCAN analyses. The location of the visualised MNI axial slices is denoted by the blue lines on the sagittal slice for each comparison.

**Supplementary Table 1:** Detailed statistical results yielded by SCCAN lesion mapping analyses. Subtests with <15 persistently impaired patients are presented in grey. Group denotes analysis timepoint (acute/persistent), N denotes the number of patients included in each analysis, range statistic reports the model weight of significant voxels, and Nsig (all) denotes the total number of significant voxels (with both positive and negative model weights). Nsig (>0) denotes the number of voxels with positive model weights, indicating that damage to these voxels is associated with worse behavioural performance. Clusters reports the number of significant voxel clusters, sparseness reports the optimal sparseness identified in the optimisation procedure, and CV P reports the P-value of the cross-validation analyses.

|  | Group | N | Range Stat | Nsig (all) | Nsig (>0) | Clusters | Sparseness | CV P |
| --- | --- | --- | --- | --- | --- | --- | --- | --- |
| **Executive** |  |  |  |  |  |  |  |  |
| Trail Making |  |  |  |  |  |  |  |  |
|  | Acute | 382 | 0 - 0 | 0 | 0 | 0 | < -0.001 | 0.535 |
|  | Persistent | 337 | 0 - 0 | 0 | 0 | 0 | < -0.001 | 0.682 |
| **Language** |  |  |  |  |  |  |  |  |
| Picture Naming |  |  |  |  |  |  |  |  |
|  | Acute | 406 | 0 - 1 | 7218 | 7218 | 6 | 0.041 | 0.039 |
|  | Persistent | 404 | 0 - 0 | 0 | 0 | 0 | 0.05 | 0.182 |
| Semantics |  |  |  |  |  |  |  |  |
|  | Acute |  |  |  |  |  |  |  |
|  | Persistent | 270 | 0 - 0 | 0 | 0 | 0 | -0.035 | 0.49 |
| Sentence Reading |  |  |  |  |  |  |  |  |
|  | Acute | 400 | -1 - 0.97 | 120172 |  | 6 | -0.301 | < 0.001 |
|  | Persistent | 394 | 0 - 1 | 75054 | 75054 | 2 | -0.201 | < 0.001 |
| **Memory** |  |  |  |  |  |  |  |  |
| Orientation |  |  |  |  |  |  |  |  |
|  | Acute | 407 | 0 - 0 | 0 | 0 | 0 | 0.006 | 0.273 |
|  | Persistent | 403 | 0 - 0 | 0 | 0 | 0 | 0.563 | 0.38 |
| Verbal Recall |  |  |  |  |  |  |  |  |
|  | Acute | 402 | -0.55 - 1 | 82836 |  | 6 | 0.35 | < 0.001 |
|  | Persistent | 395 | 0 - 0 | 0 | 0 | 0 | -0.01 | 0.328 |
| Episodic Recognition |  |  |  |  |  |  |  |  |
|  | Acute | 379 | 0 - 1 | 29145 | 29145 | 2 | 0.099 | 0.004 |
|  | Persistent | 367 | 0 - 0 | 0 | 0 | 0 | < 0.001 | 0.108 |
| **Number** |  |  |  |  |  |  |  |  |
| Number Writing |  |  |  |  |  |  |  |  |
|  | Acute | 407 | -0.63 - 1 | 172662 |  | 6 | -0.383 | < 0.001 |
|  | Persistent | 400 | 0 - 1 | 15672 | 15672 | 2 | 0.06 | < 0.001 |
| Calculation |  |  |  |  |  |  |  |  |
|  | Acute | 407 | -0.53 - 1 | 1690 |  | 3 | -0.009 | 0.005 |
|  | Persistent | 400 | 0 - 0 | 0 | 0 | 0 | < -0.001 | 0.297 |
| **Praxis** |  |  |  |  |  |  |  |  |
| Gesture Imitation |  |  |  |  |  |  |  |  |
|  | Acute | 401 | -0.98 - 1 | 149759 |  | 33 | -0.451 | 0.006 |
|  | Persistent | 378 | 0 - 0 | 0 | 0 | 0 | < -0.001 | 0.247 |
| **Visuospatial** |  |  |  |  |  |  |  |  |
| Left Egocentric Neglect |  |  |  |  |  |  |  |  |
|  | Acute | 377 | -0.43 - 1 | 31404 |  | 8 | -0.136 | 0.012 |
|  | Persistent |  |  |  |  |  |  |  |
| Right Egocentric Neglect |  |  |  |  |  |  |  |  |
|  | Acute | 377 | 0 - 0 | 0 | 0 | 0 | < -0.001 | 0.217 |
|  | Persistent | 369 | 0 - 0 | 0 | 0 | 0 | 0.031 | 0.067 |
| Left Allocentric Neglect |  |  |  |  |  |  |  |  |
|  | Acute | 377 | -0.85 - 1 | 59088 |  | 24 | -0.328 | 0.003 |
|  | Persistent | 369 | 0 - 0 | 0 | 0 | 0 | 0.122 | 0.315 |
| Right Allocentric Neglect |  |  |  |  |  |  |  |  |
|  | Acute | 377 | 0 - 0 | 0 | 0 | 0 | < -0.001 | 0.151 |
|  | Persistent | 369 | 0 - 0 | 0 | 0 | 0 | -0.02 | 0.577 |

SCCAN involves a rigorous and conservative cross-validation procedure which requires a large number of both spared and impaired patients to yield significant results.^78^ It is possible that the comparatively small number of patients with persistent impairment lead these analyses to be underpowered.^84^ Alternatively, it is possible that these null results indicate that factors outside of lesion location are responsible for driving differential recovery trajectories. However, the correlates of acute impairment identified by SCCAN analyses align well with those reported in previous studies. ^14,51,52^

**Supplementary Table 2:**
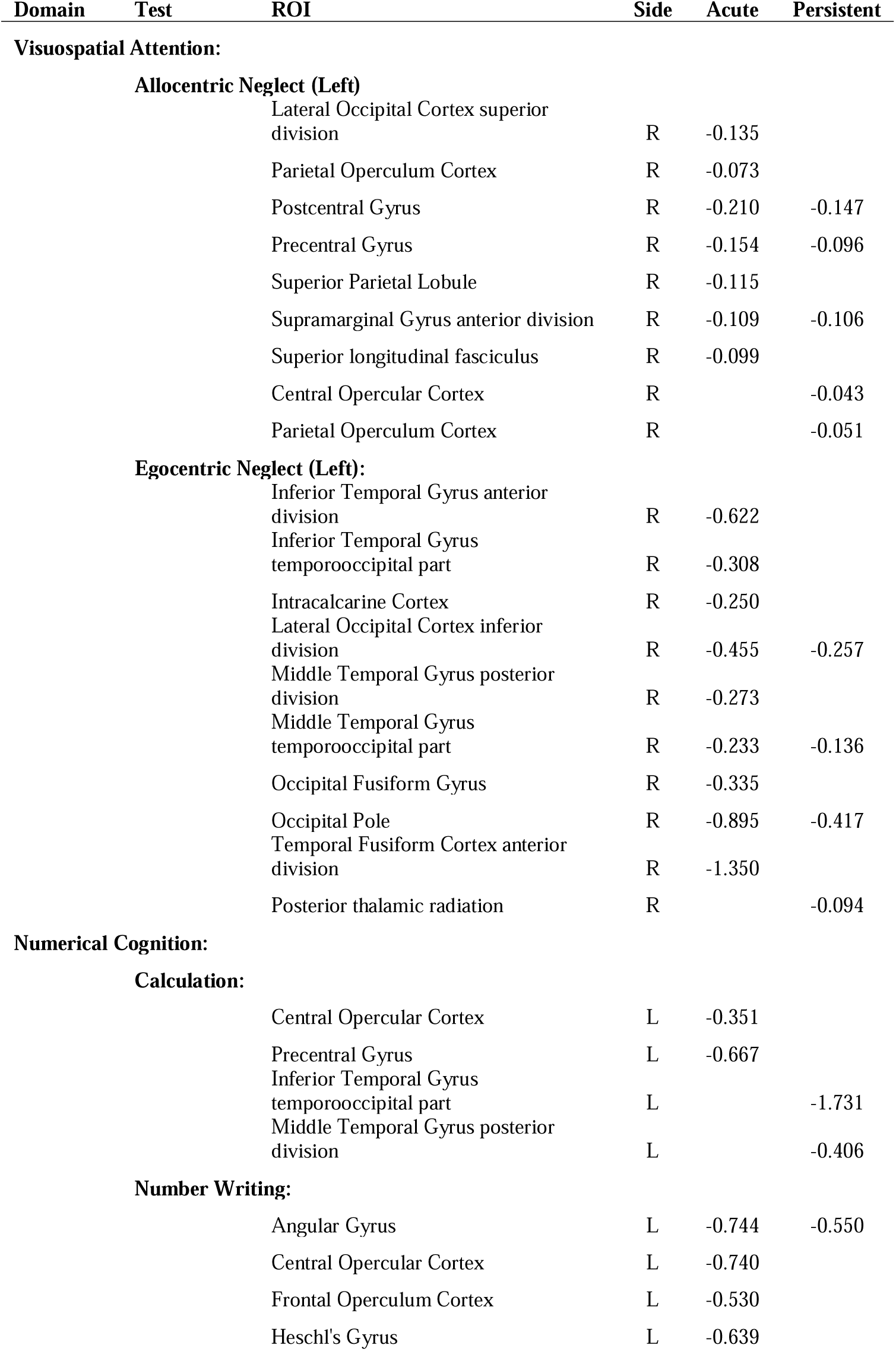
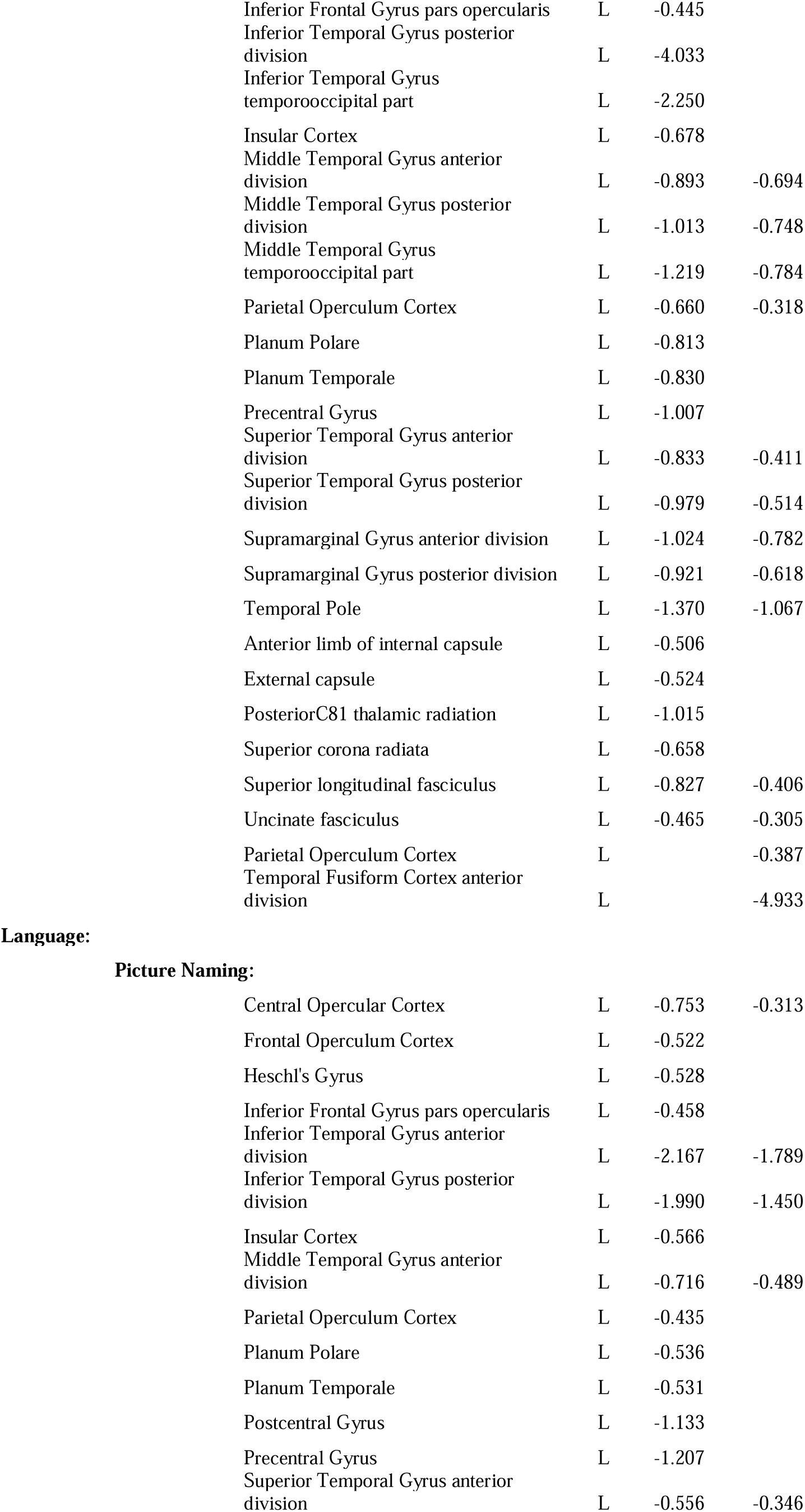
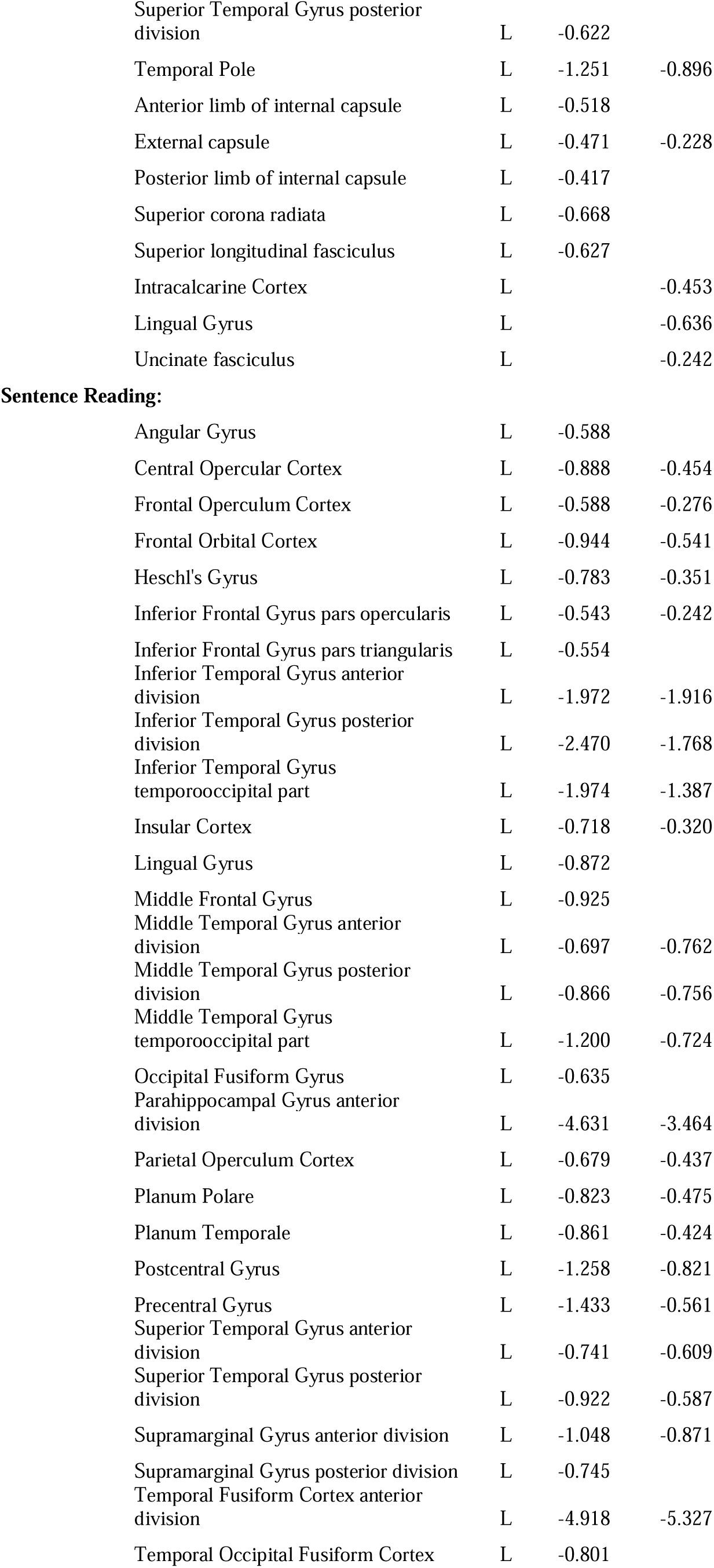
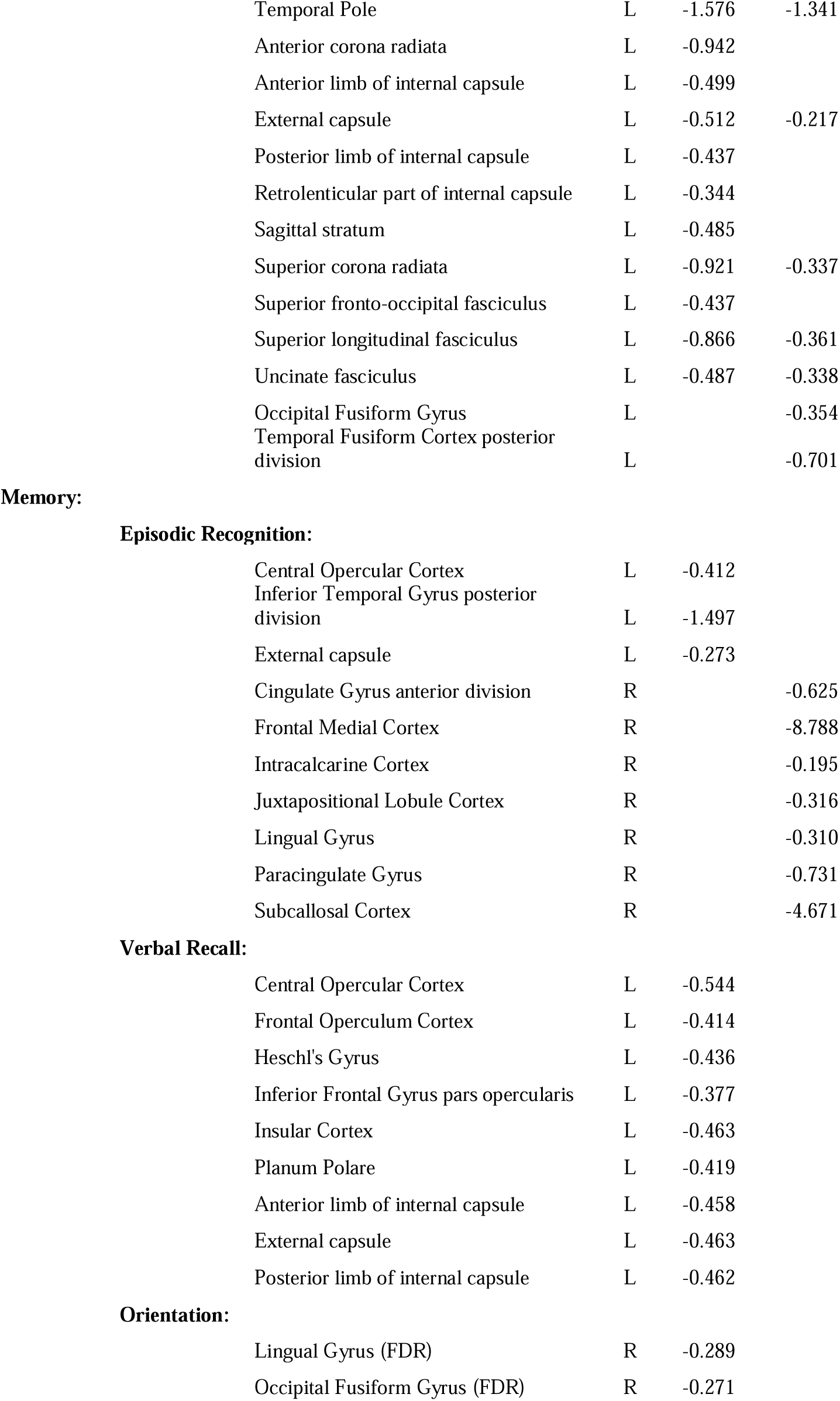
Anatomical descriptives for all significant ROI-level lesion mapping results. Domain reports the cognitive domain while Test denotes the relevant OCS subtest. Side represents the lateralisation of the significant ROI, and Acute/Persistent report the B value of the relevant ROI at each timepoint respectively. All listed ROIs (with the exception of those denoted FDR) survived Bonferroni corrections for multiple comparisons.

**Supplementary Table 3:**
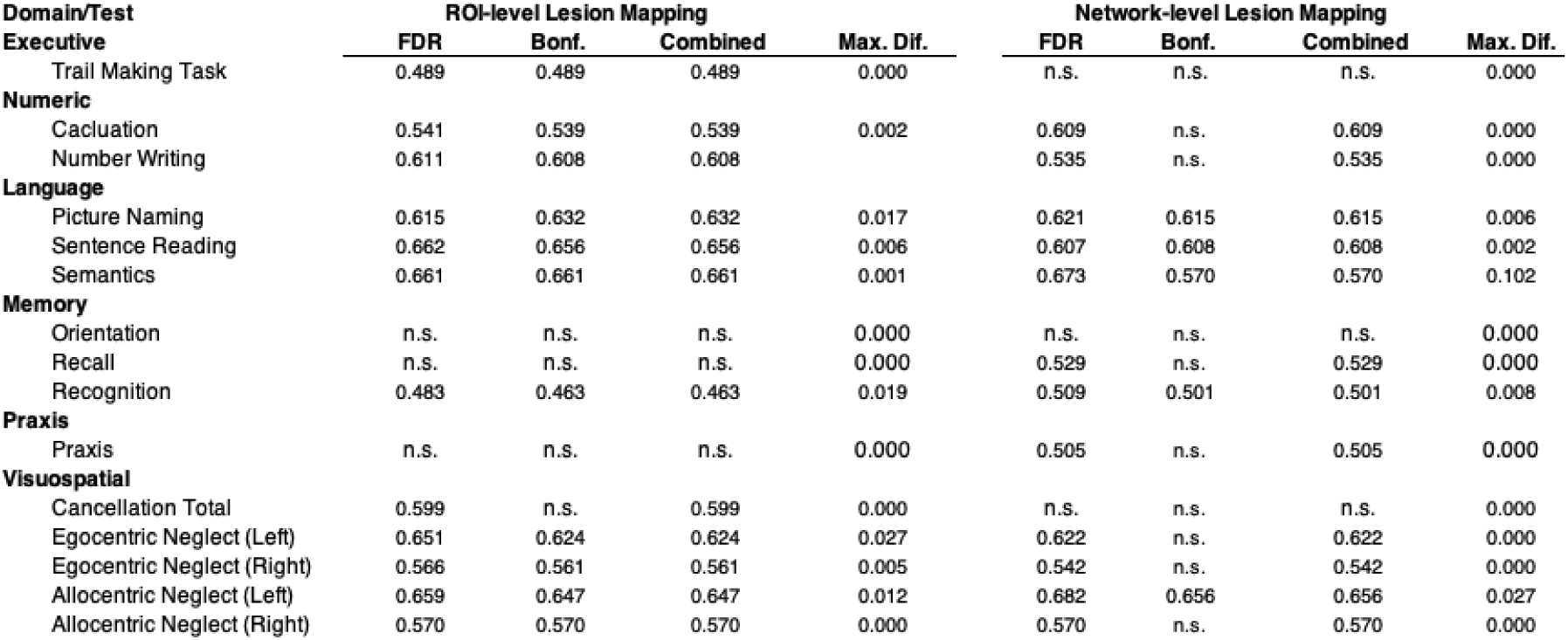
A comparison of AUC values generated by using FDR, Bonferroni, or the combined correction method used in this study (Bonferroni when significant, FDR when Bonferroni not significant). AUC values were comparable across all correction approaches. The mean difference between the highest and lowest possible AUC values was 0.008 (range = 0.00 – 0.102). These results indicate that alternate lesion mapping correction methods would not significantly modulate the resultant diagnostic model accuracy. SCCAN results are excluded from these tables as this method does not employ corrections for multiple comparisons.

